# Physical Activity and Academic Achievement among Omdurman Islamic University Medical Students 2021

**DOI:** 10.1101/2022.12.12.22283383

**Authors:** Mohammed Ahmed Abdelrahman, Alaa Abuelgasim, Hiba Allah Hassan Marouf, Abrar Mohamed Alalim

## Abstract

**Background:** Physical activity is important for the prevention of non-communicable diseases and for growth, development and general health.

there is evidence that has emerged from university studies suggesting, that physical activity may have a positive effect on student’s academic achievement.

**Objectives:** The aim of this study is to assess the relationship between physical activity and academic achievement among Omdurman Islamic University Medical Students in 2021.

**Materials and methods:** A des sectional University based study done on faculty of medicine in Omdurman Islamic university. The sample of 295 medical students was taken from the first, second and third year in the Faculty of Medicine in Omdurman Islamic University who studied during the academic years of 2018-2019. The students are between the ages of 18-23 years old and are physically healthy (both active and inactive candidates).A systematic random technique sample was taken from the students during the year 2020.Data Collected through a structured interviewer-administered questionnaire The data was coded and analyzed by a statistician using Statistical Package for Social Science (SPSS) version 23.

**Results:** shows that most students (56.3%) were physically active and the most types of physical activity practiced by the physically active students were walking (42.8%) and play football (25.9%)), and the barriers of being physically active among those physically inactive students were the study load in most of the students (22.4%). the results indicate that there is no direct association between physical activity and academic achievement (P value= o.7).

**Conclusion:** Many factors overlap in affecting the academic achievement of students, some of them have a great effect, while student’s physical activity level appears have a lesser effect.

**Recommendations:** Expanding the study to include a number of universities and majors with a recommendation for Omdurman Islamic University to include the importance of physical activity in its curricula.

**Conflict of interest:** No conflict of interest.

## 1. Introduction

Physical activity is important for the prevention of no communicable diseases and for growth, development and general health. ^(1)^ The World Health Organization recognizes that the minimum physical activity that helps prevent chronic diseases is 150 minutes of moderate physical exercise or 75 minutes of high intensity physical activity per week or both. ^(1)^ Walking is considered as the easiest and safest way to exercise by rate of 10,000 steps/day. ^(1)^

Physical activity has a major role in promoting people’s health, and its significance increased in developed and developing countries. ^(2)^ In 2002, the World Health Organization celebrated World Health Day under the theme of moving for health, focusing on physical activity. ^(2)^ Studies from 142 countries (93% of the world’s population) showed that physical inactivity leads to 6.4% of deaths around the world related to heart disease, strokes, diabetes, in addition to colon and breast cancer. ^(2)^ The World Health Organization believes that 25% of the aforementioned diseases can be prevented by encouraging physical activity, with an aim to reduce incidence to 10% by 2030. ^(2)^

Academic achievement is the extent to which a learner achieves the goals of an educational institution, whether they are long or short-term goals. A learner must have a final status such as a high school diploma and a university bachelor’s degree, and most institutes measure academic achievement by the grade point average (GPA).^(10)^ The importance of academic achievement emerges from providing information that assesses strengths and weaknesses of the educational system. This will help in building educational policies, improving strengths and addressing shortcomings. Some studies indicate that physical activity has an effect on academic achievement and classroom behavior, but there is debate on the extent of this effect and there are a number of elements associated with physical activity that must be examined such as type, duration, frequency, and whether increase in physical activity over the recommended are negatively affect academic achievement. ^(8)(9)^

There is an urgent need to introduce physical activity within students’ hours because of prevalence of non-communicable diseases. ^(6)^

It is difficult to determine the variables that affect the relationship between physical activity and academic achievement, such as attitude toward the physical activity. ^(3-4)^ The link between physical activity and academic achievement is generally unclear but there are findings that suggest a noteworthy influence that physical activity may have effect on the performance of underachieving students. ^(5)^

### 1.1. Statement of the problem

Academic achievement is of great importance in the student’s academic career and also contributes to his competition later in the labor market. A number of factors interfere in influencing academic achievement and any factors that positively affect academic achievement are considered of great importance, so this study was conducted to study the effect of physical activity on academic achievement because the situation among Sudanese university not raised before.

### 1.2. Justification (Rationale)

There is a lack of awareness about the importance of physical, mental and social health ^(17)^. A minority of the population are physically active and understand the significant effect it may have on their health.

Being physically active is important especially when linked with academic achievement which is fundamental for student promotion and general development. There is a need to conduct more studies in this area, in order to achieve the highest quality of education as an end result and due to the importance of academic achievement to the students’ career and in labor market.

### 1.3 Study objectives

#### 1.3.1. General objectives

To assess the relationship between physical activity and academic achievement among Omdurman Islamic University Medical Students in 2021.

#### 1.3.2. Specific Objectives

1. To assess the type, duration and frequency of physical activity of a study group.
2. To determine the academic achievement level of the study group.
3. To identify the most common barriers associated with physical activity among the study group.
4. To determine the association between physical activity and academic achievement.

### 2 Literature Review

Physical activity has an impact on the social, academic and economic life of individuals, and the methods of physical activity vary from light to heavy. Its importance in students’ lives is evident in its effect on their academic achievement, their desire to learn, their level of participation and interaction, along with a number of other factors involved.

#### 2.1. Physical activity

is defined by the World Health Organization as “any physical movement produced by the skeletal muscles of a person that requires the consumption of energy” ^(7)^. And by this, it includes whenever a person does a sport in his spare time or when he moves to work as an example or part of the work he performs ^(7)^.And it has been proven that moderate-intensity aerobic physical activity and vigorous-intensity aerobic physical activity, improves one’s health ^(7)^. The most common forms of physical activity are walking, cycling, sports, football, etc. Studies have also shown that physical activity has a number of health benefits as it helps prevent non-communicable diseases such as heart disease and cancer, as well as improves mental and psychological health, reduces depression and anxiety, and increases learning and thinking skills ^(7)^. .In adults, physical activity helps maintain physical fitness (cardiorespiratory and muscular fitness), cardiovascular health (blood pressure, dyslipidemia, glucose, and insulin resistance) as well as bone health, as it increases cognitive outcomes (academic performance, executive function) and contributes significantly ^(7)^. It has also been found to maintain mental health (reduced symptoms of depression) and helps reduce obesity ^(7)^.

According to the recommendation of the World Health Organization, people within the age group of university students, should engage in regular physical activity whether moderate intensity for 150-300 minutes per week, or vigorous intensity physical activity for 75-150 minutes per week ^(11)^. Globally, 1 in 4 adults does not meet the recommended international levels ^(11)^. With physical activity, we can avoid 5 million deaths annually at a global scale if we are more active. Inactive people have an increased risk of dying by 20% to 30% compared to the active people ^(10)^. It has also been found that more than 80% of adolescents worldwide, suffer from the effects of low Physical Activity. ^(11)^

In low-income countries, 12% of men and 24% of women were not sufficiently physically active as opposed to 26% of men and 35% of women in high-income countries ^(11)^. This lack of physical activity is attributed to the sedentary behavior of people at work and at home, plus increased use of transportation. ^(11)^

Physical inactivity also has an underlying effect on the economy as it increases the cost of health care progressively and reduces production ^(11)^. According to 2016 estimates, physical inactivity has cost the global health system about 54 billion US dollars and led to economic losses estimated at about 14 billion US dollars ^(11)^. It is also estimated that between 1-3% of the health system expenditures are due to physical inactivity ^(11)^.

#### 2.2. Academic achievement

“describes academic results that indicate the extent to which the student has achieved his own learning goals and the institution’s achievement of long or short-term educational goals, and thus it refers to the completion of educational degrees such as a bachelor’s or diploma”. Academic achievement is often measured through examinations and continuous evaluation of the student ^(12)^.

Educational institutions define the educational and cognitive goals that the student must acquire, including knowledge acquisition in addition to understanding in a specific field of thought such as science, health and history ^(12)^. So academic achievement is a multi-faceted structure that includes different areas of learning and the definition of academic achievement depends on the indicators used to measure it, which are general indicators such as knowledge acquired from the educational system, or indicators based on school curricula, such as academic GPA, or cumulative indicators of academic achievement such as academic certificates ^(12)^. And academic achievement has an important role in the lives of individuals in developed countries and measured by the GPA, which is based on the educational grades obtained by the student which in turn has an impact on his or her career in the future ^(12)^.

Many factors overlap in influencing the student’s academic achievement. However, the most important of which are social and economic factors such as parental support, standard of living and the family’s financial situation ^(15)^. Studies have shown that students who fall under the lower socio-economic class are the ones with the lowest academic achievement ^(15)^. Also, environmental factors have an impact on academic achievement, such as the geographical location of the educational institution and the number of students ^(15)^. In addition, psychological factors have an impact on academic achievement such as problems of learning difficulty, or psychological and social integration ^(15)^. Some educational institutions also select students based on previous academic achievement and their distinction, which has an impact on their future achievement ^(15)^.

A number of researchers divided the factors affecting academic achievement, especially among medical students, into two parts ^(16)^. The first is internal factors, such as the student’s competence in the English language, classroom schedules, class size and environment, complexity of the scientific material, the role of professors in the educational process and the extent of technology use in study and exams. As for the external factors, they are outside the educational institution, such as family, financial and social problems, along with his/her practice of physical activity, sports, etc., as well as good communication skills ^(16)^. All of the factors mentioned above have a role in improving academic achievement ^(16)^.

There is a further study on providing Sedentary Adults with Choices for Meeting Their Walking Goals. This study was designed to test different ways of meeting the new CDC recommendations for physical activity stating that all Americans at least 2 years of age should obtain 30 minutes of moderate intensity activity on most days of the week ^(1)^. Results showed that all groups significantly (P ≤ 0.05) improved their aerobic fitness, their systolic blood pressure and increased their physical activity at the end of the program ^(1)^.

#### 2.3. Previous studies

In further research about the relative influence of individual, social and physical environment determinants of physical activity, it was shown that environmental determinants of health are receiving growing attention in the literature, although there is little empirical research in this area ^(4)^. The study was conducted to examine the relative influence of individual, social environmental and physical environmental determinants of recreational physical activity ^(4)^. It involved a community survey of 1803 healthy workers and home-makers aged 18-59 years living in metropolitan Perth, Western Australia ^(4)^. Overall, 59% of respondents exercised as recommended. Recreational facilities located near the home were used by more respondents than facilities located elsewhere ^(4)^. The most frequently used facilities were informal: the streets (45.6%); public open space (28.8%) and the beach (22.7%) ^(4)^. the results suggest that access to a supportive physical environment is necessary, but may be insufficient to increase recommended levels of physical activity in the community ^(4)^. Given the popularity of walking in the community, it is recommended that greater emphasis be placed on creating streetscapes that enhance walking for recreation and transport ^(4)^.

An additional study was carried out on low levels of physical activity in Sudanese individuals with some features of metabolic syndrome: it is a population based study ^(17)^. The aim of this study was to evaluate the level of physical activity among Sudanese population ^(17)^. A descriptive cross sectional study composed of 323 participants from Khartoum state, Sudan. Data was collected using pretested designed questionnaires based on previously validated Global Physical Activity Questionnaire ^(17)^. The demographic and physical measurement includes blood glucose, anthropometric and blood pressure ^(17)^. Results showed that males were 59.9%, females 47.1% and the prevalence of inactivity was 53.8% ^(17)^. Despite the fact that males are more active than females (P value Conclusion that Physical activity in Sudanese women was significantly decreased in comparison with men) ^(17)^.

According to statistics, more than a quarter of the world’s adult population (about 1.4 billion adults) do not engage in sufficient physical activity ^(7)^. Just 1 in only 3 women and 1 in 4 men do not engage in the minimum level of physical activity recommended to maintain good health levels of physical inactivity are twice as high in high-income countries compared to low-income countries ^(7)^. Statistics obtained in the year 2016 show that 28% of adults aged 18 years and over (men 23% and women 32%) do not meet the global recommendation of at least 150 minutes of moderate activity per week or 75 minutes of vigorous activity per week ^(7)^.

A study conducted aims: to explore physical activity (PA) habits among the medical students and examine the correlation with their grade point average (GPA) achievement at the College of Medicine, King Saud University, Riyadh, Saudi Arabia. A cross-sectional study was conducted among the medical students (n = 409), during the academic year 2012–2013 ^(19)^. Students’ physical activity habits were self-reported. GPA was collected and analyzed with SPSS software. Results showed that out of 409 students, 193 (47.2%) students reported being physically active ^(19)^. There result showed a significant positive association between students’ PA habits and high-GPA achievement (p = 0.001) ^(19)^. The greatest odds ratio of high GPA was found among the fourth year students (OR = 3.08, p = 0.025) and fifth year students (OR = 5.07, p = 0.010). In addition, a significant association was found between the normal BMI and high-GPA achievers (p = 0.016) ^(19)^.However, no statistically, significant association was found between body fat percent (BF %) and GPA ^(19)^.

Another study aimed to explore the prevalence of physical activity among medical and health sciences students at Cyberjaya University College of Medical Sciences and to determine the relationship of their physical activity level with their academic achievement and self-determination level ^(20)^. An analytical cross-sectional study was conducted among 244 Medical and Health Sciences undergraduate students at CUCMS from January to April 2017 using a self-administered short-form version of the International Physical Activity Questionnaire (IPAQ-SF) and the third version of the Behavioral Regulation in Exercise Questionnaire (BREQ-3) ^(20)^. Multiple regression models were fitted using SPSS version 20 to examine the relationships between the study variables ^(20)^. Results showed that half of the male students (51.7%) were in the health-enhancing physical activity (HEPA) group, as compared to only 24.7% of females ^(20)^. The odds of having a good grade point average was twice as high among HEPA active students (odds ratio [OR] = 1.89, 95% CI [1.09, 3.27], P = 0.023) than among non-HEPA active students ^(20)^. Furthermore, the odds of being HEPA active was higher for males (OR = 3.16, 95% CI [1.61, 6.14], P < 0.01) than for females and higher for overweight students than for normal weight students (OR = 2.58, 95% CI [1.24, 5.57], P = 0.017) ^(20)^. The odds of being HEPA active was 1.79 times higher for each unit increase in the integrated regulation score (OR = 1.79, 95% CI [1.14, 2.91], P = 0.020) ^(19)^. Conclusion: The prevalence of physical inactivity was higher among females than males ^(20)^. This study also confirmed a significant association between physical activity level and academic achievement. HEPA active students performed better academically than those who were non-HEPA active ^(20)^.

Studies indicate that there is a close relationship between physical activity and academic achievement and that the relationship between them is positive in most cases despite the presence of a number of other factors that positively affect students’ academic achievement, including internal factors related to the study environment and teaching methods as well as other external factors related to the student’s mental, financial and social abilities.

### 3. Methodology

#### 3.1. Design of the study

Descriptive cross-sectional University based study.

#### 3.2. Study Setting

The study setting is the faculty of medicine in Omdurman Islamic university, which lies in the southern side of Omdurman-Sudan –Abu Sead area.

Omdurman Islamic university was chosen as the area in which this research take place due to its accessibility and feasibility.

#### 3.3. Study Population

The study population is a sample of medical students of the Faculty of Medicine in Omdurman Islamic University ages of 18-23 years (1130 students as obtained from the academic affairs office of the university in 2018) studying in the academic year of 2018-2019. The students are physically healthy (both active and inactive candidates). Excluded candidates are those outside the average age of university students (between 18 and 23 years), those physically unhealthy, and candidates with social factors as parents education and home environment that may affect their academic achievement.

#### 3.4. Sample Size

For the sample to be scientific and reflect the characteristics of the population, a sample of 295 students who studying at first, second and third years was taken, calculated from the 1130 of the whole population by this formula:

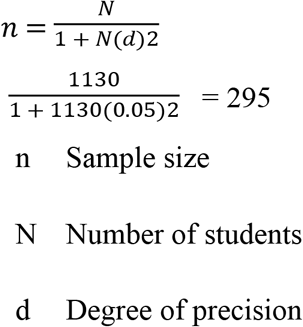

With a confidence level of 95%, confidence interval of 9 and margin of error 5%.

#### 3.5. Sampling Technique

A systematic random technique sample was taken from the medical students of the faculty of medicine in Omdurman Islamic university during the year 2021 and the sample size was proportionally distributed to the 1^st^, 2^nd^ and 3^rd^ years. If individual refused to participate we took the next candidate with the same interval and then return to the original interval.

#### 3.6. Data Collection Methods and Tools

A structured interviewer-administered questionnaire was prepared containing 3 sections. The first section consists of the socio-demographic variables such as age, sex and origin. The second section was contain the type of physical activity, duration and its frequency. The third section was contain the academic achievement of students, such as poor, average or excellent, others factors affect Academic Achievement rather than Physical Activity, and the GPA was obtained. The data was obtained from the academic affairs office of the university. The data collected by telephone interview.

##### Study Variables

###### 3.6.1. Dependent variables: academic achievement

###### 3.6.2. Independent variables: physical activity (frequency-duration)

###### 3.6.3. Background variables

Demographic data (age - gender) - origin (central Sudan (which includes states of Khartoum, Gezira, and White Nile), North of Sudan (which include Northern and River Nile states), West of the Sudan (which include Darfur and Kordofan States), East of the Sudan (which include Red see, Kasala and Gadaref States) south of the Sudan (Blue Nile, South Kordofan and Senar States)), finical status, life style, definition of physical activity.

#### 3.7. Data Management and Analysis

The data was coded and analyzed by a statistician using Statistical Package for Social Science (SPSS) version 23.The descriptive data was presented as graphs and charts.

The association between categorical variables was calculated to determine significance level (p value) accepted when it is less than 0.05 by using descriptive comparative and correlation analysis and leaner regression analysis which was done by statistician.

#### 3.8. Ethical Considerations

The ethical clearance taken from the institutional review board at The International University of Africa.

Permission was granted by the faculty of medicine authorities in Omdurman Islamic University.

Written informed consent before starting interviewing the students. Confidentiality was preserved.

## 4. Results

**Table No (1):** the relationship between academic achievement and the Status of Physical Activity among Omdurman Islamic University Medical Students-2021.

| Are you physically active? | N | Mean GPA | Std. Deviation |
| --- | --- | --- | --- |
| Yes | 166 | 3.09 | .47 |
| No | 129 | 3.16 | .44 |

**Table No (2):** cross tabulation of Physical Activity in Relation to Student Origin among Omdurman Islamic University Medical Students (Males-Females) 2021.

|  |  |  | Are you physically active? |  | Total |
| --- | --- | --- | --- | --- | --- |
|  |  |  | Yes | No |  |
| Origin | Central of Sudan | Count | 108 | 80 | 188 |
|  |  | % within Are you physically active? | 65.1% | 62.0% | 63.7% |
|  | West of Sudan | Count | 13 | 12 | 25 |
|  |  | % within Are you physically active? | 7.8% | 9.3% | 8.5% |
|  | East of Sudan | Count | 8 | 4 | 12 |
|  |  | % within Are you physically active? | 4.8% | 3.1% | 4.1% |
|  | North of Sudan | Count | 32 | 25 | 57 |
|  |  | % within Are you physically active? | 19.3% | 19.4% | 19.3% |
|  | South of Sudan | Count | 5 | 8 | 13 |
|  |  | % within Are you physically active? | 3.0% | 6.2% | 4.4% |
| Total |  | Count | 166 | 129 | 295 |
|  |  | % within Are you physically active? | 100.0% | 100.0% | 100.0% |

Table (2) shows that there is no association between student origin and their ability to be physically active (P value= 0.6).

**Table No (3):** the relationship between academic achievement and the frequency of Physical Activity among Omdurman Islamic University Medical Students-2021.

|  | N | Mean GPA | Std. Deviation |
| --- | --- | --- | --- |
| daily | 85 | 3.12 | .51 |
| weekly | 77 | 3.06 | .43 |
| monthly | 4 | 3.17 | .14 |
| Total | 166 | 3.09 | .47 |

**Table No (4):** the relationship between academic achievement and the type of Physical Activity among Omdurman Islamic University Medical Students-2021.

|  | N | Mean GPA | Std. Deviation |
| --- | --- | --- | --- |
| Walking | 69 | 3.18 | .46 |
| Running | 17 | 2.95 | .58 |
| Football | 46 | 3.07 | .47 |
| other | 34 | 3.05 | .41 |
| Total | 166 | 3.09 | .47 |

**Table 5.** Table No (5): the relationship between academic achievement and the parents’ education among Omdurman Islamic University Medical Students-2021.

|  | N | Mean | Std. Deviation |
| --- | --- | --- | --- |
| Yes | 274 | 3.09 | .48 |
| NO | 4 | 3.07 | .28 |
| One of them | 17 | 3.27 | .32 |
| Total | 295 | 3.09 | .47 |

## 5. Discussion

The purpose of this study is to assess the relationship between physical activity and academic achievement.

The study shows that most of the participants were males with mean age of (21.13 ± 1.42) and most of are living in their house (with their families). The main origin of students from central Sudan (which includes states of Khartoum, Gezira, and White Nile). (Figures 1, 2 & 3)

**Figure No (1):**
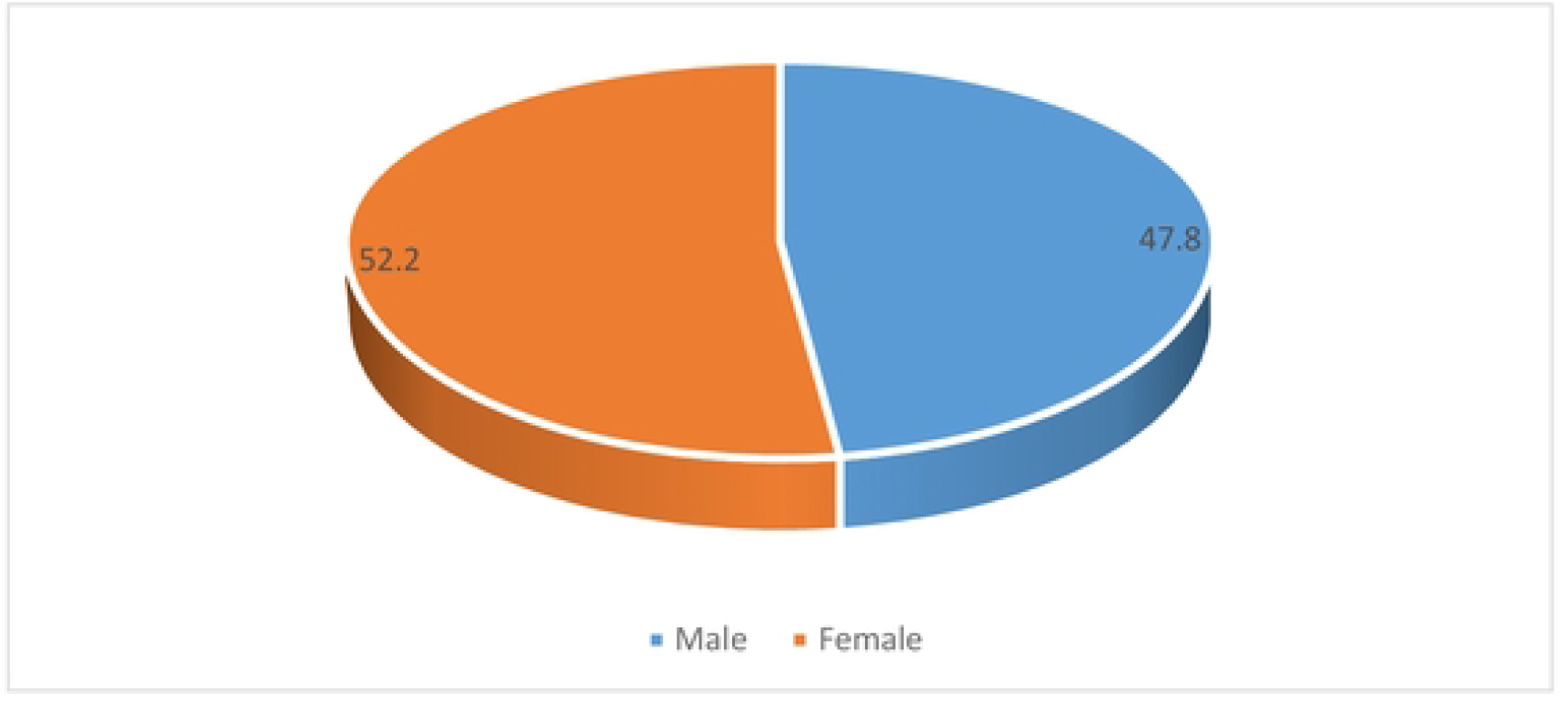
gender of Omdurman lslamic University Medical Students-2021. Figure (1) shows that 52.2 were females and 47.8 were males. Text NO (1): The mean age of Omdurman Islamic University Medical Students (21.1 3 ± 1.42).

**Figure No (2):**
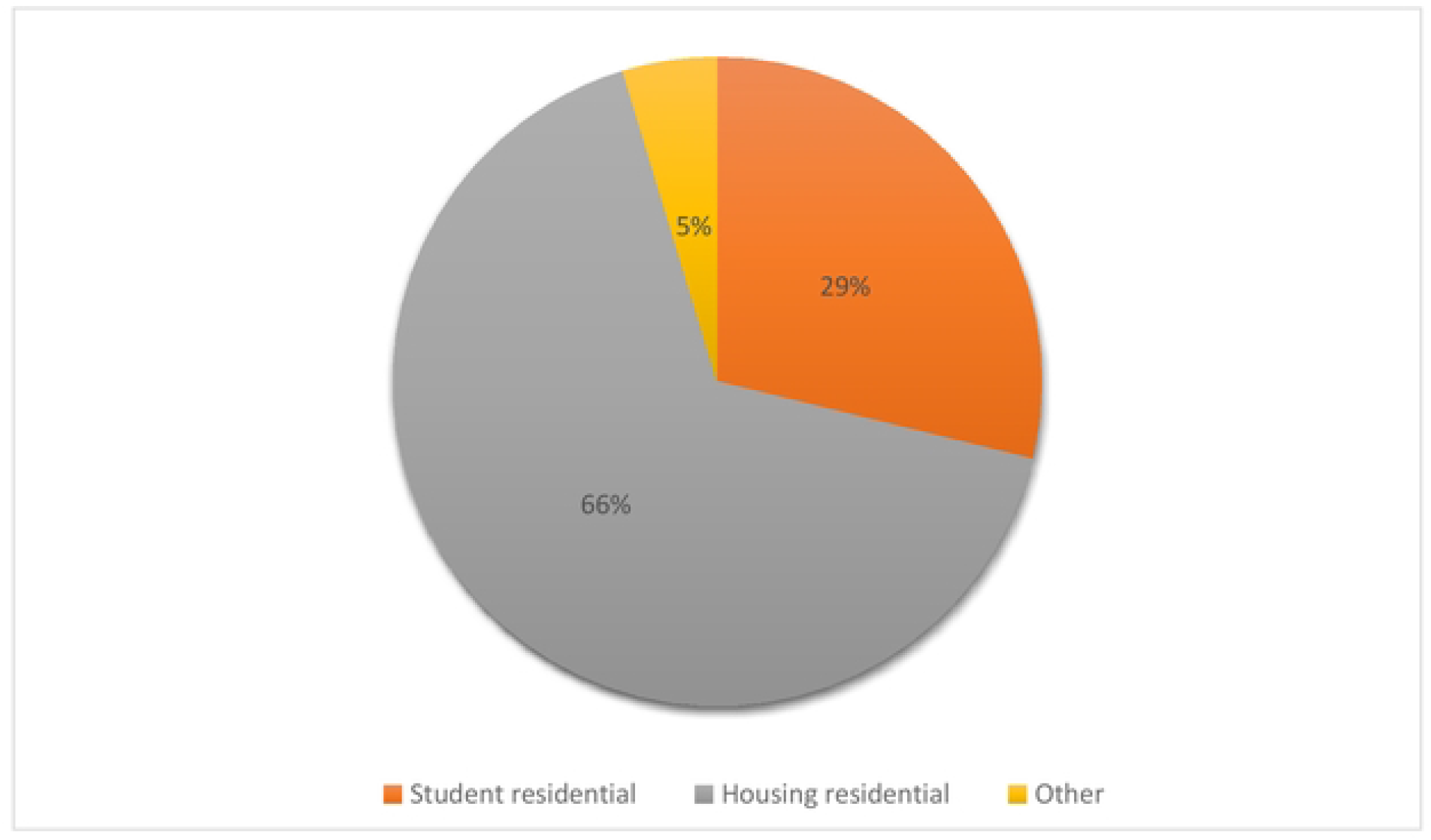
residential status of Omduraman Islamic University Medical Students-2021. Figure (2) shows that Most of the students are living in housing (with their families) residential (66.4%),while the student residential were only (28.8%) and the rest of the students (4.7) were living with their friends.

**Figure No (3):**
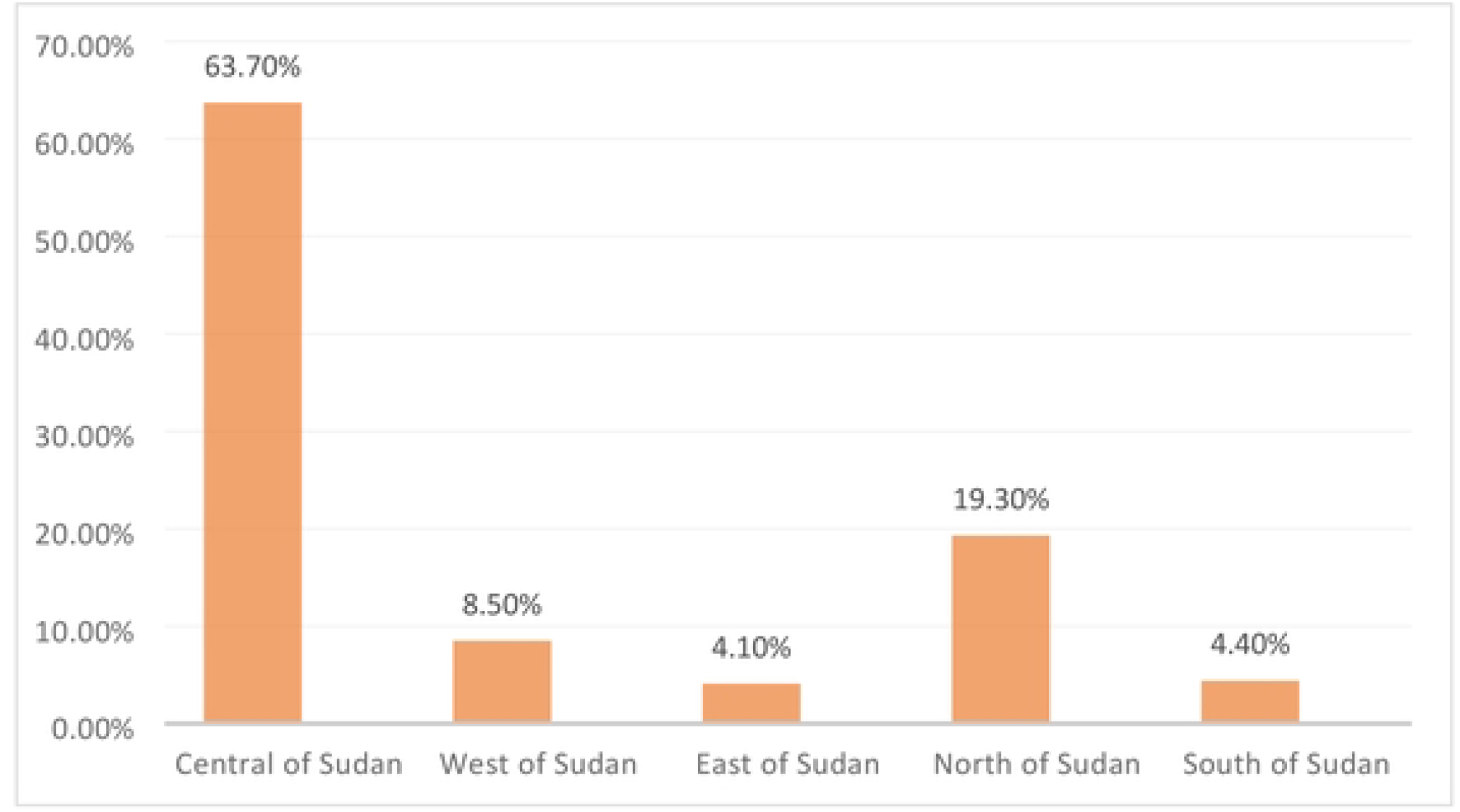
origin of Omduraman Islamic University Medical Students-2021. Figure (3) shows that most of students (63.7%) were originally from central Sudan (which includes states of Khartoum, Gezira, and White Nile).

The results also shows that most of the students’ parents were educated and the educational level of most students’ parents was university level and the students had a GPA mean of 3.08 ± .47 with a P value of o.5. this finding reveals, that there is no significant relationship between parents education and AA which is dissimilar with the result from previous studies done by (Abdulghani HM, and et al) and (Niromand E, and et al) that proved the direct impact of the parental level of education as the main drivers for increasing academic achievement ^(13)(14)^. (Figure 4 & table 5)

**Figure No (4):**
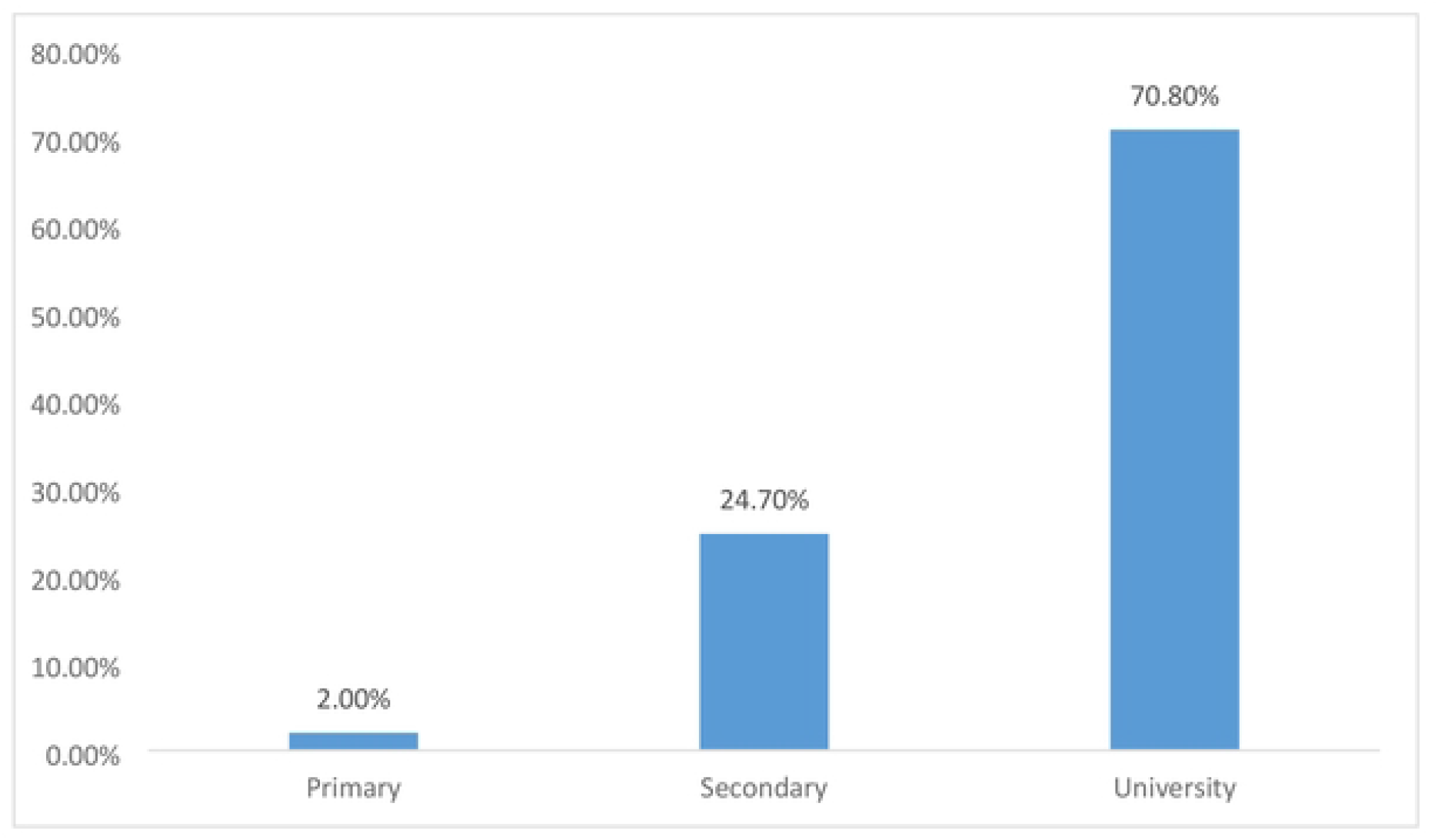
educational level of parents among Omdurman Islamic University Medical Students-2021. Figure (4) shows that the educational level of students’ parents was primary school (2%), secondary school (24.7%) and university level (70.8%).

The results show that the level of knowledge about physical activity among students according to the World Health Organization’s definition of physical activity was high, which agreed with result from Queens University ^(21)^. This is can be due to their perception about physical activity from their academic background of medicine. (Figure 5)

**Figure No (5):**
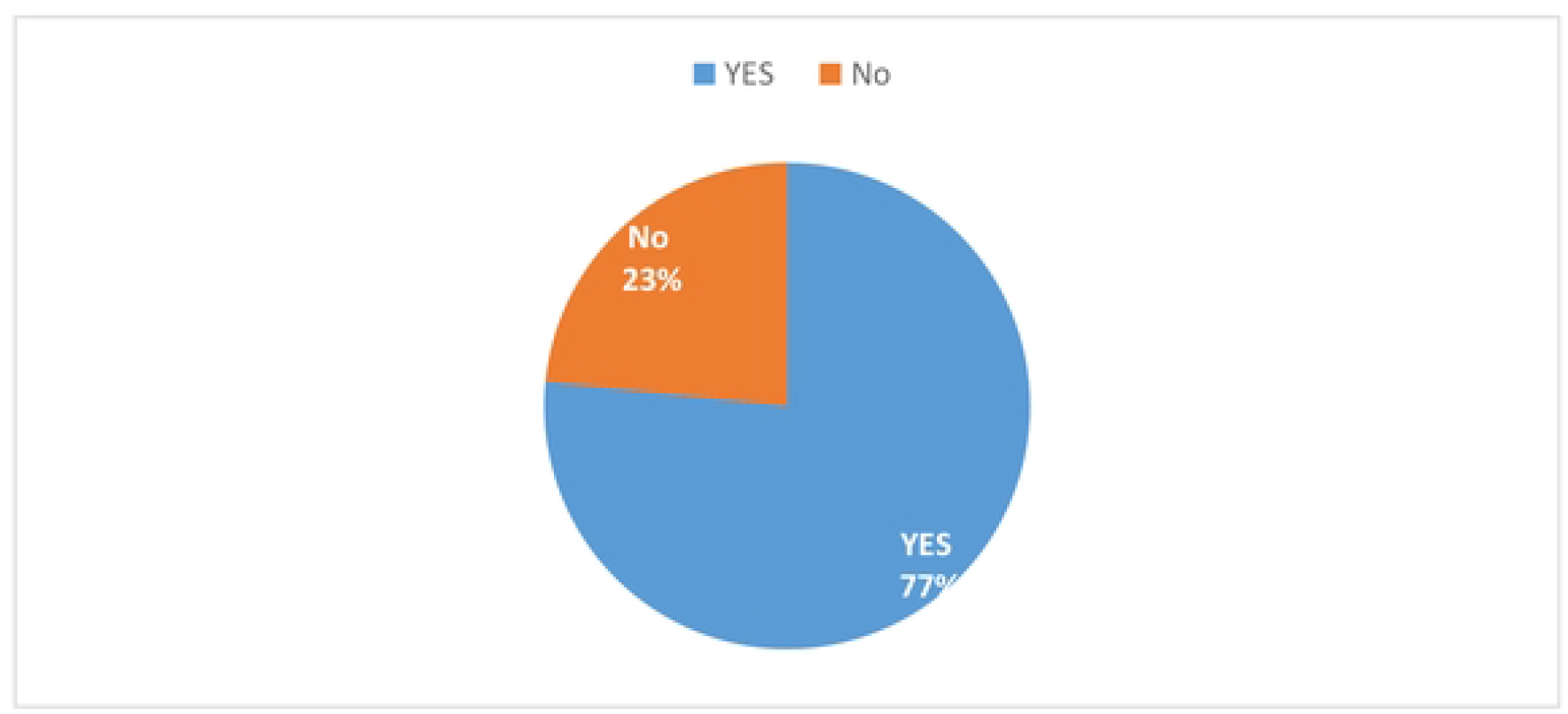
Knowledge about physical activity among Omdurman Islamic University Medical Students-2021. Figure (5) Shows that most of students (76.6%).have a good knowledge about physical activity according to the World Health Organization’s definition of physical activity.

The results shows that most of students were physically active which agreed with result from King Saud University ^(11)^. Most of the physical active students were males which is agreed with the previous study done in Khartoum-Sudan by Khalil S, and et al^(6)^ and with other a study done in Malaysia revealed the contrary, where males are physically more active than females ^(20)^, in spite of cultural differences and high number of female in the population (Figure 5).

The most common barriers among those physically inactive students were the study load. This back to the field of study of those student which is medicine which contain a large amount of information with restricted time. (Figure 6)

**Figure No (6):**
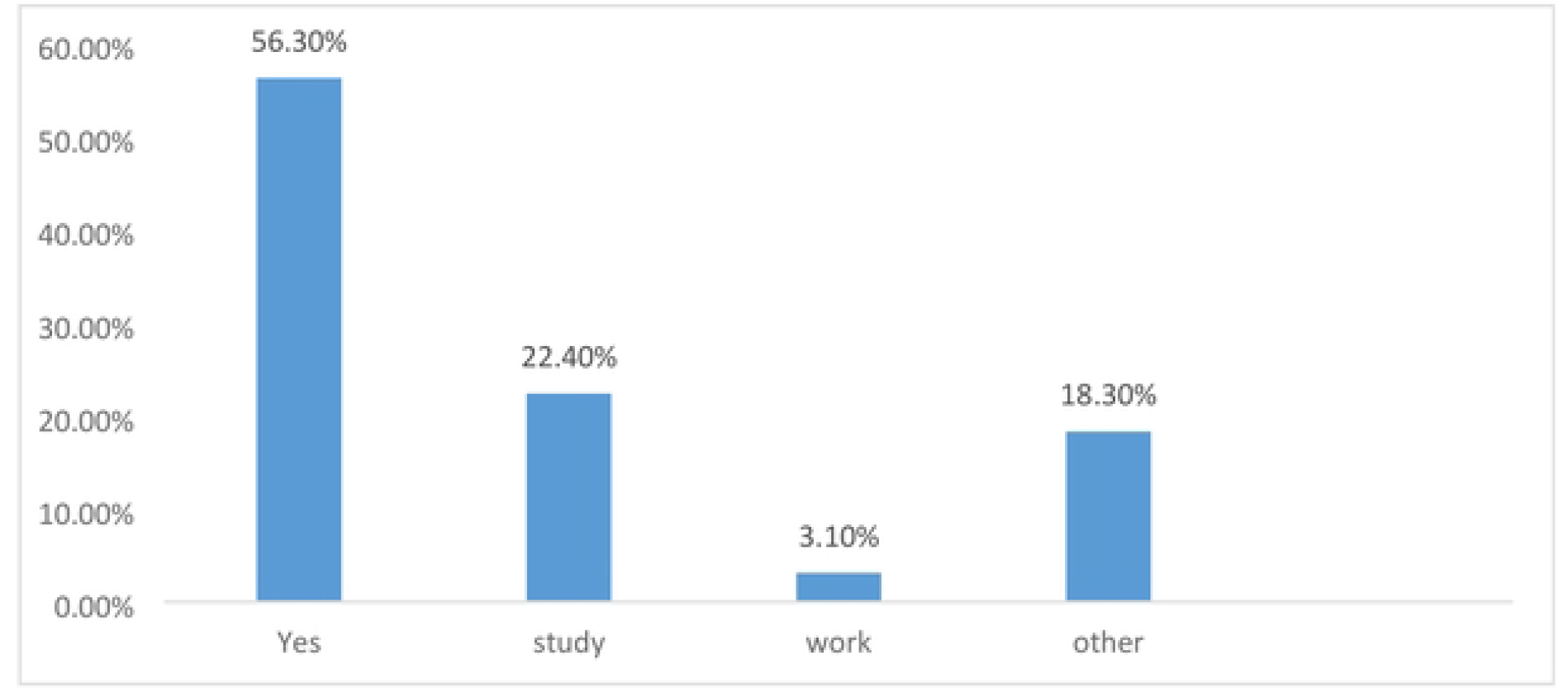
status of physical activity and its barriers among Omdurman Islamic Univerity Medical Students -2021. Figure (6) shows that most of students (56.3%) were physically active and the barriers of being physically active among those physically inactive students were the study load in (22.4%) in most of the students.

Walking is the main type of physical activity practiced by those physically active students. This is maybe due to the wide surface area of the university campus which provides space for students to walk, play football, and run; while the rest prefer the gym in case of males and Zumba for females. Most of physically active students practice physical activity on a dilly base. (Figures 7&8)

**Figure No (7):**
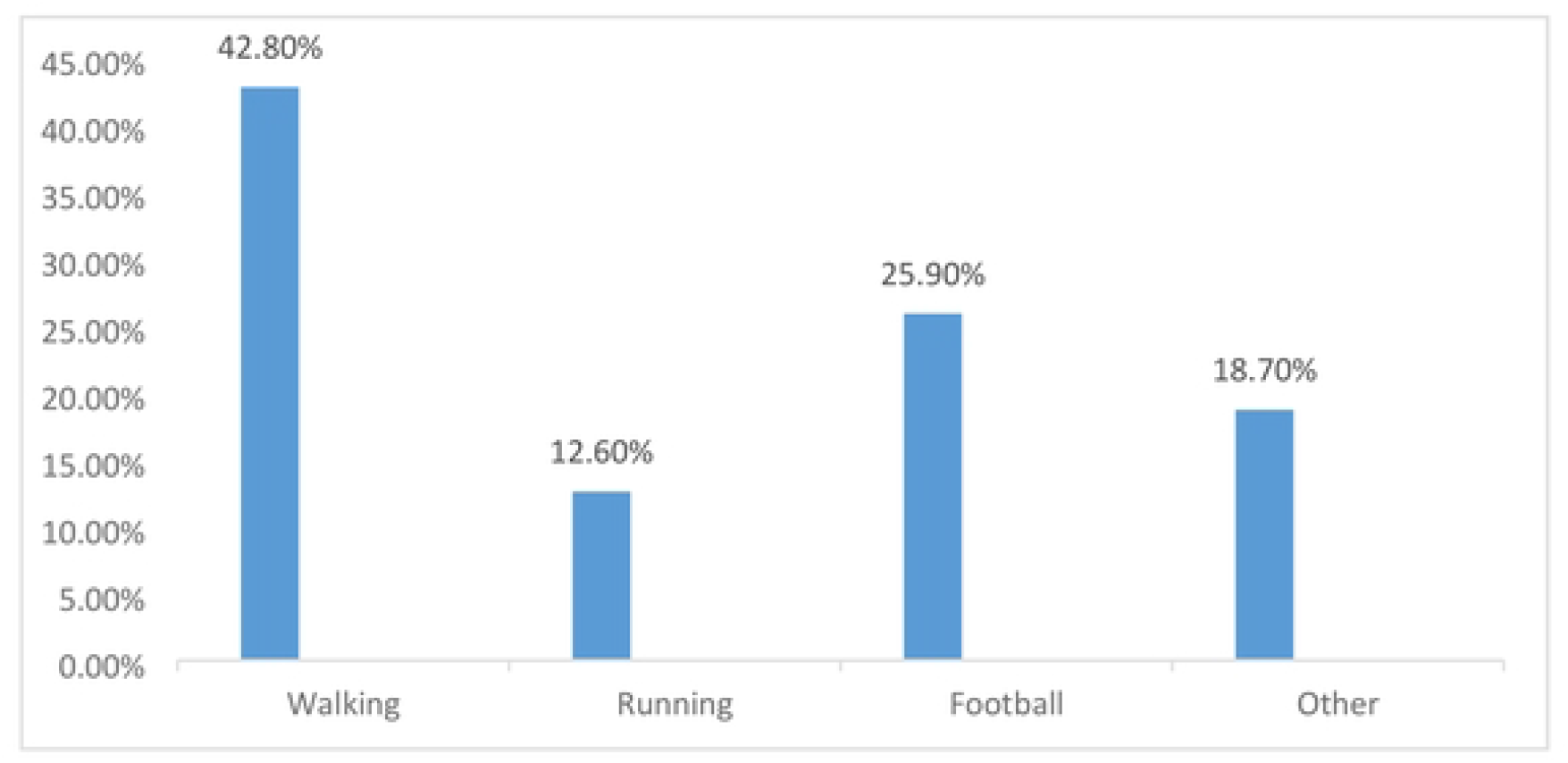
types of physical activity done by Omdurman Islamic University Medical Students-2021. Figure (7) shows that the most types of physical activity practiced by the physically active students were walking (42.8%) and playing football (25.9%).

**Figure No (8):**
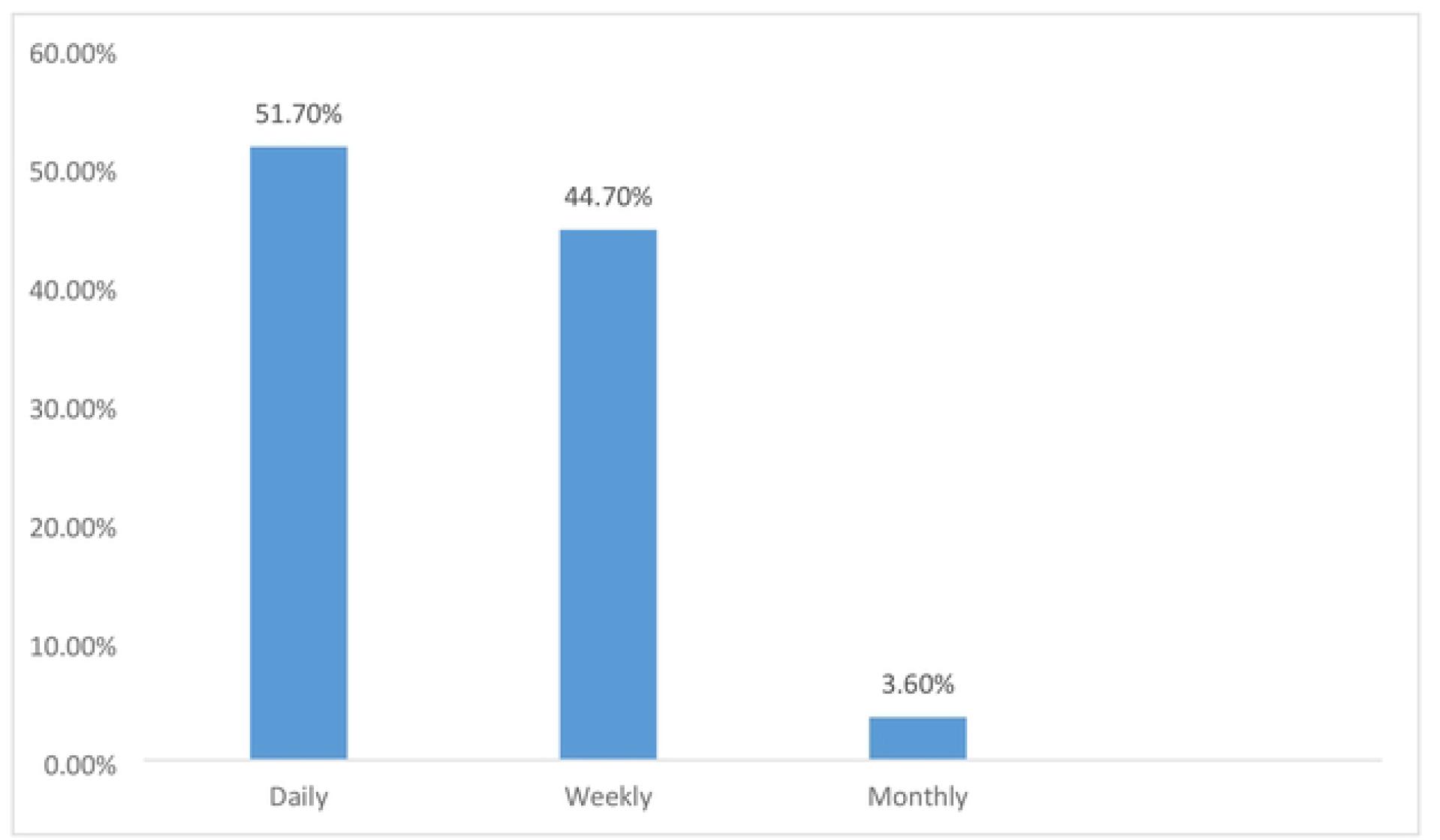
Frequency of physical activity done by Omdurman lslamic University Medical Students-2021. Figure (8) shows the frequency of physical activity for students were daily in (51.7%), weekly (44.7%) while only (3.6%) practiced physical activity monthly. Text no (2) The mean duration of physical activity was (116.74 ± 120.37) minutes.

The result shows that for most physically active students exercise was a routine. (Figures 9&10).

**Figure No (9):**
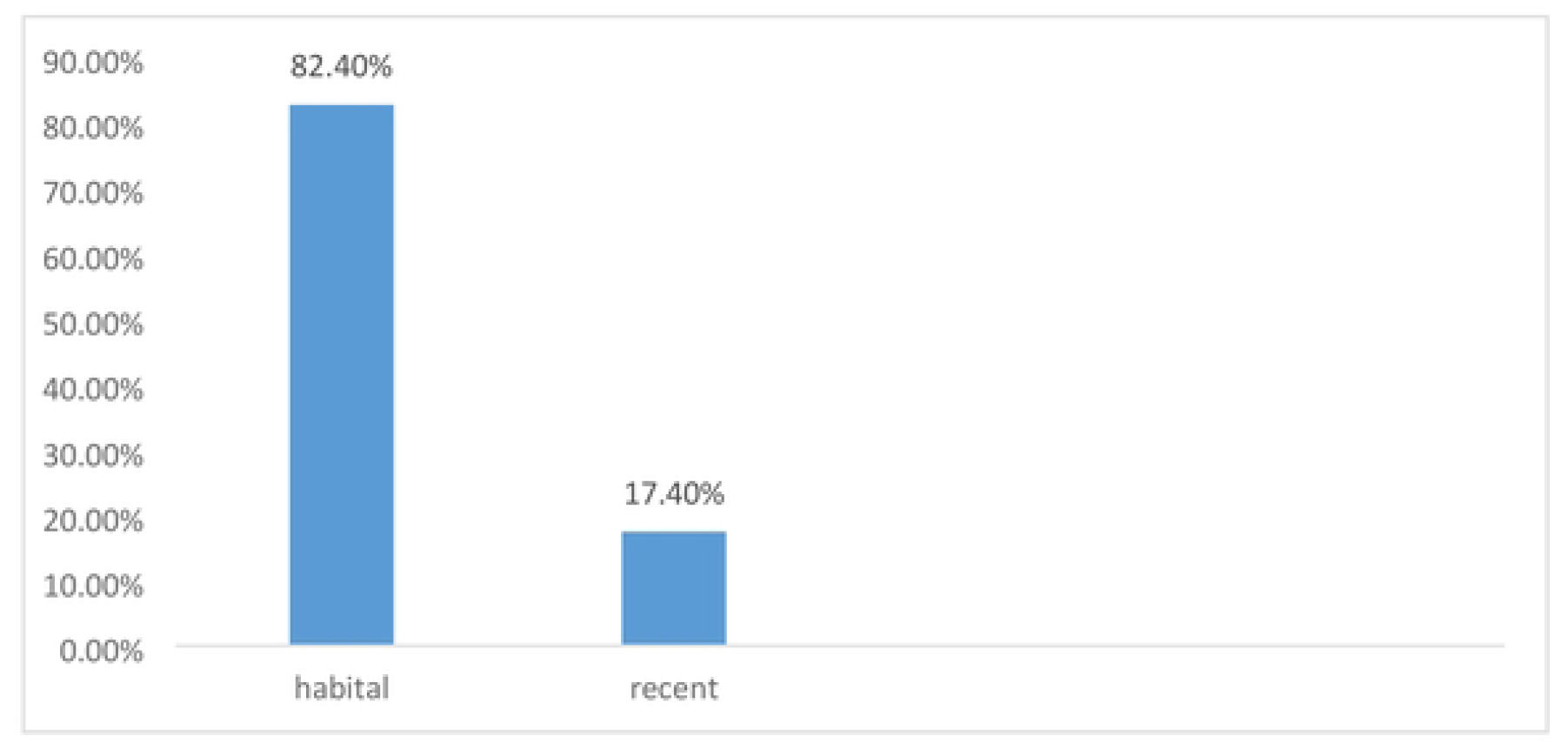
status of physical activity as a habit done by Omdurman Islamic University Medical Students-2021. Figure (9) shows that for most physically active students (83.4%), exercise was a habit while for the rest (17.4%) it was a recently added activity for them.

**Figure No (10):**
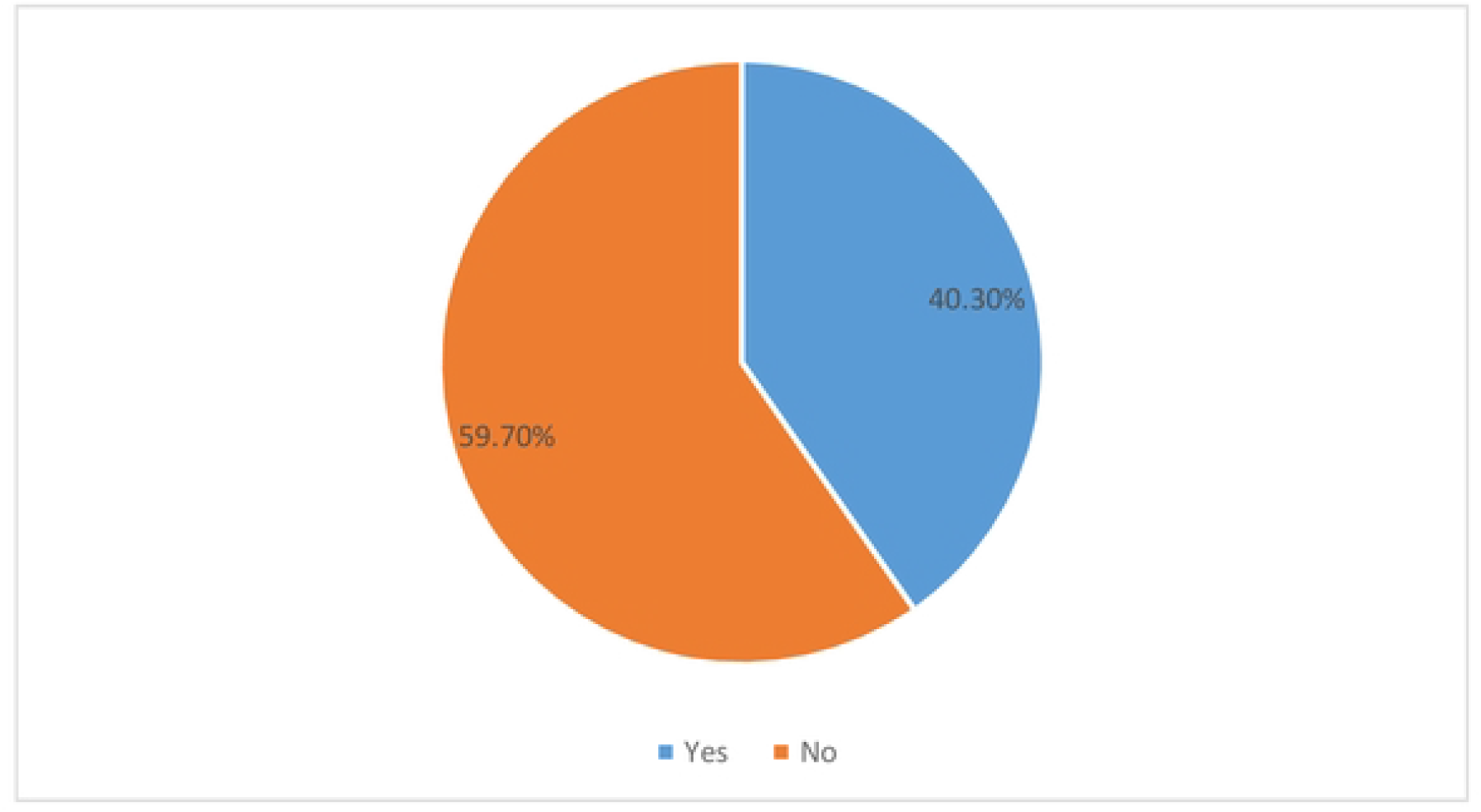
status of physical activity as a leisure done by Omdurman Islamic University Medical Students-2021. Figure (10) shows that most of the physically active students (59.7%) did not practice physical activity as a leisure while the rest (40.3%) took physical activity as a leisure

Most of students agree that physical activity has a direct effect on their academic achievement and most of them claimed that physical activity increase their academic achievement. (Figures 11&12)

**Figure No (11):**
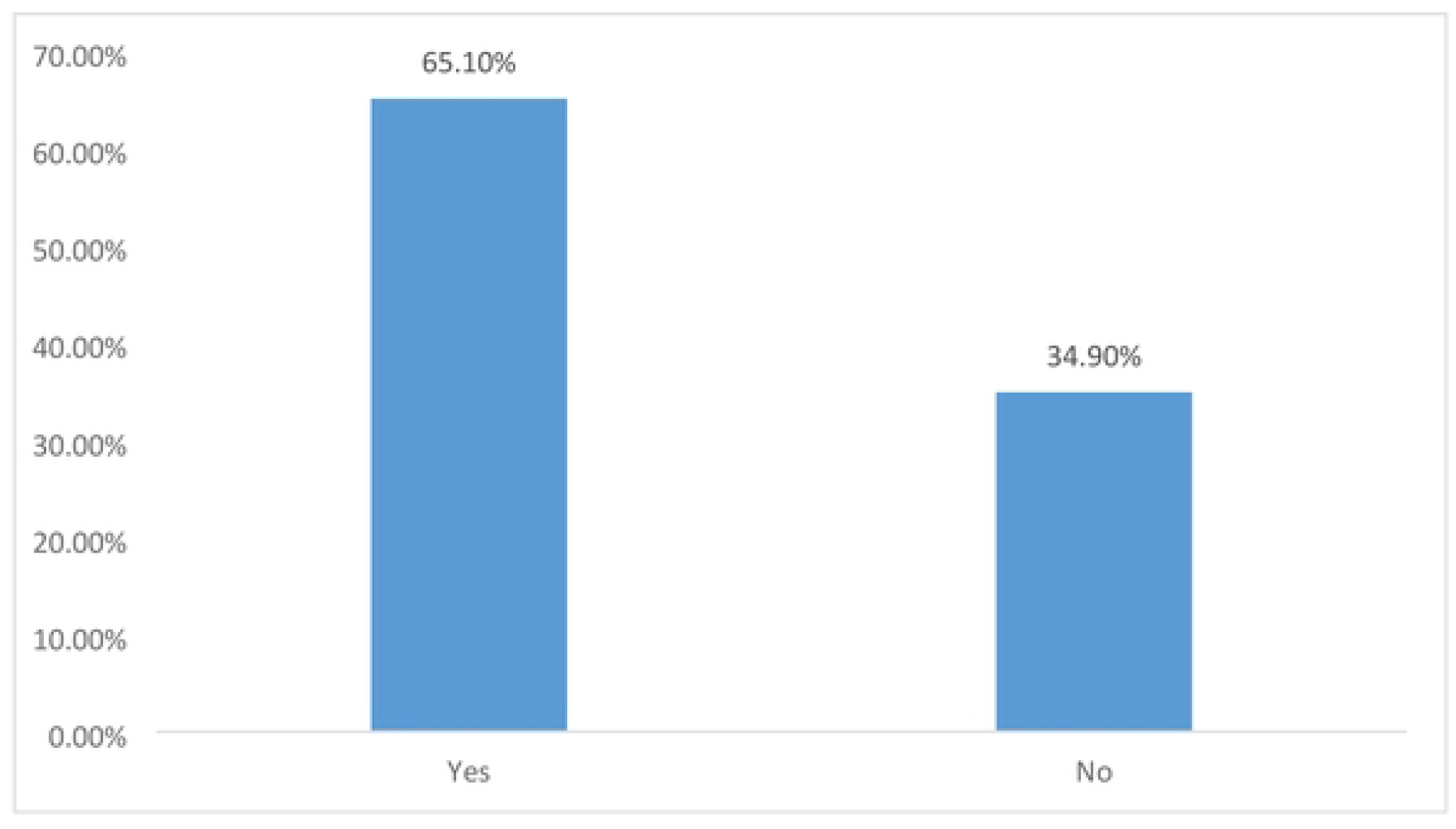
opinion of Omdurman Islamic University Medical Students on physical activity effect on their academic achievement-2021. Figure (11) shows that most of students (65.1%) agree that physical activity has a direct effect on their academic achievement, while the rest (34.9%) report that physical activity has no effect on their academic achievement

**Figure No (12):**
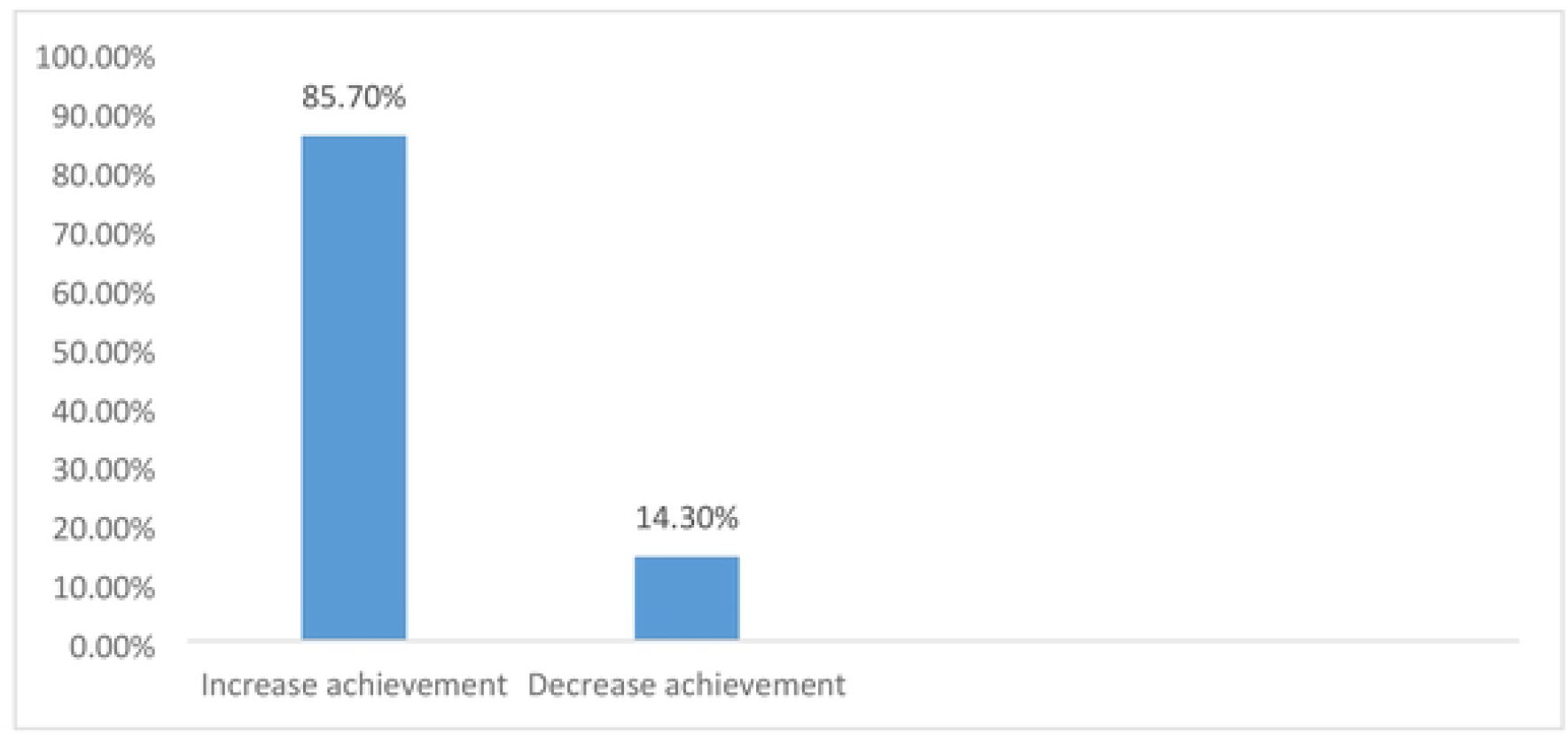
the effect of physical activity on your academic achievement among Omdurman Islamic University Medical Students-2021. Figure (12) shows that students who reported on whether physical activity affected their academic achievement claimed there is an increase in their academic achievement in (85.7%) of cases while the rest (14.3%) reported that it decreased their academic achievement.

The result shows that the mean GPA of physically active students was within the range of 3.09 ± .47, while physically inactive students had a GPA mean of 3.16 ± .44 with a P value of o.7, this finding reveals, that there is no significant relationship between PA and AA which is dissimilar with the result from King Saud University (p = 0.001) ^(11)^. And this result is due to most students reportedly living in a home environment suitable for studying and have a high interest in studying and achieving academic excellence, and reported that the university teaching methods had a positive impact on their academic achievement, and this results applies to previous studies that proved the direct impact of the home environment and student interest for academic achievement as the main motives for increasing academic achievement ^(13) (14) (15)^. (Figures 13, 14&15)

**Figure No (I3):**
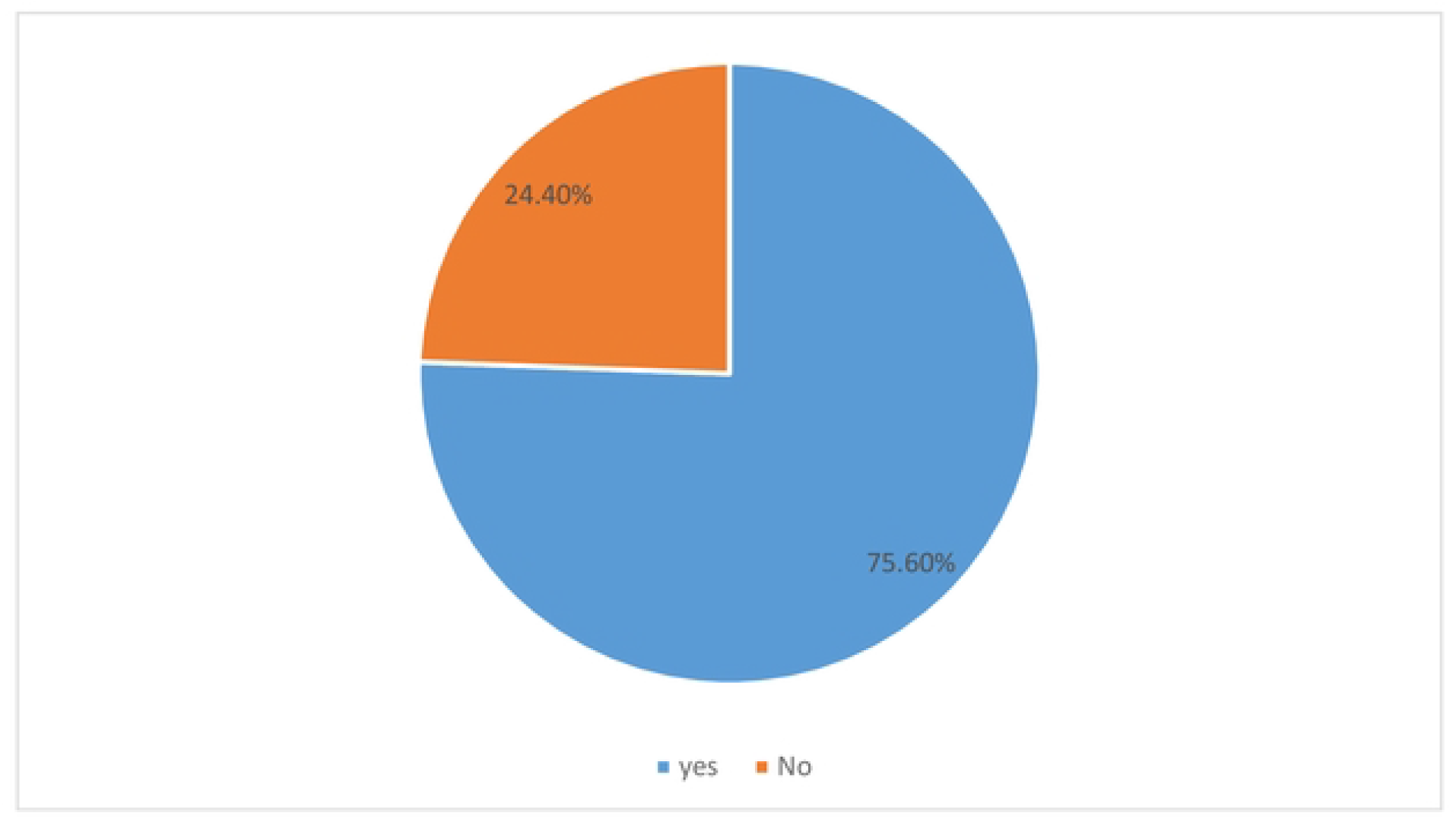
status of home environment as conducive to studying among Omdurman Islamic University Medical Students-2021. Figure (13) shows that most of students (75.6%) reportedly living in a home environment suitable for studying versus (24.4%) not living in a home environment suitable for studying.

**Figure No (14):**
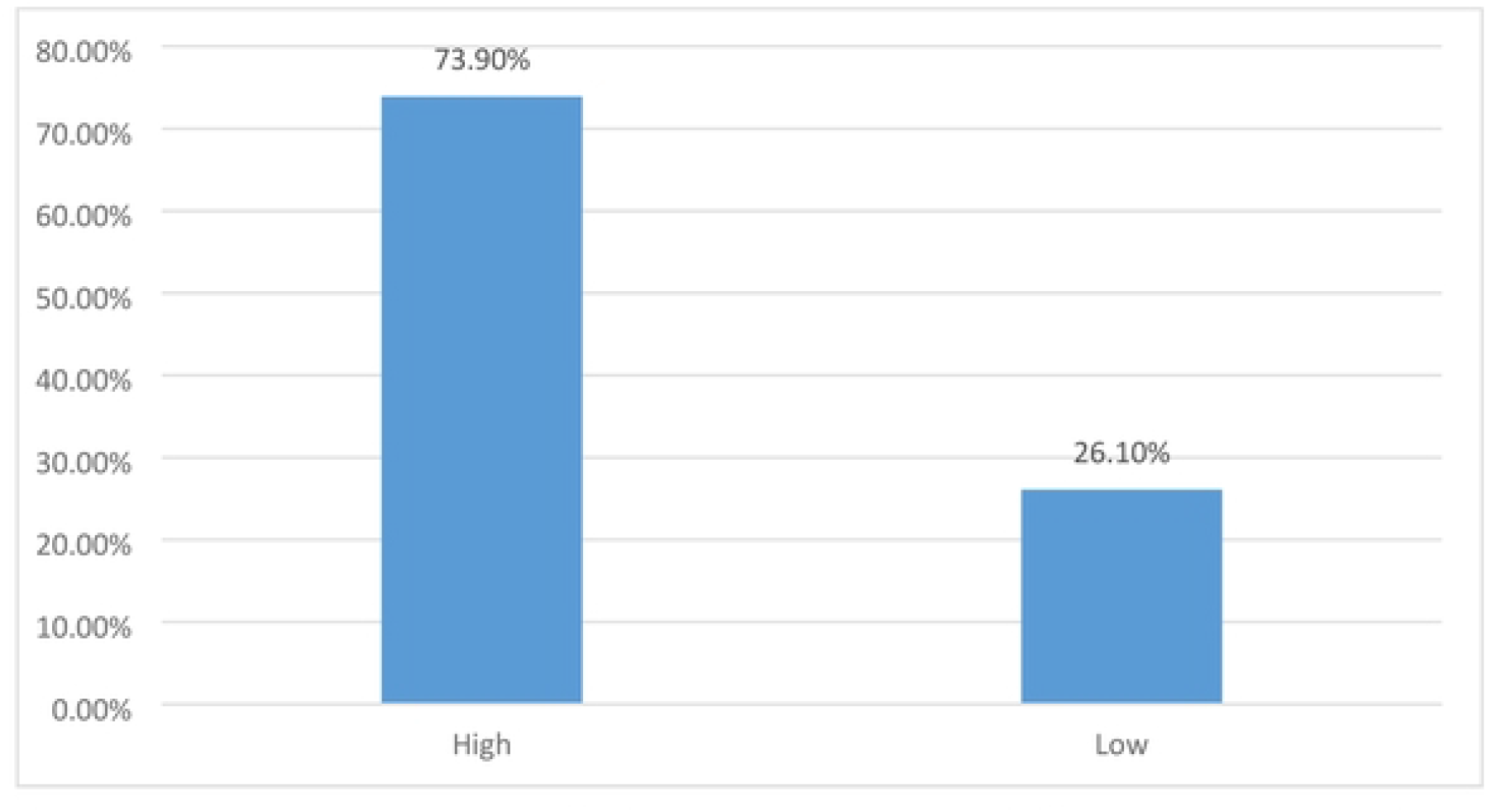
status of interest to have high academic achievement among Omdurman Islamic University Medical Students (Males-Females)2021. Figure (14) shows that most of students (73.9%) have a high interest in studying and achieving academic excellence, while (26.1 %) show low interest in studying and academic achievements. TextNO (3): The GPA mean (3.12 ± .46)

**Figure No (15):**
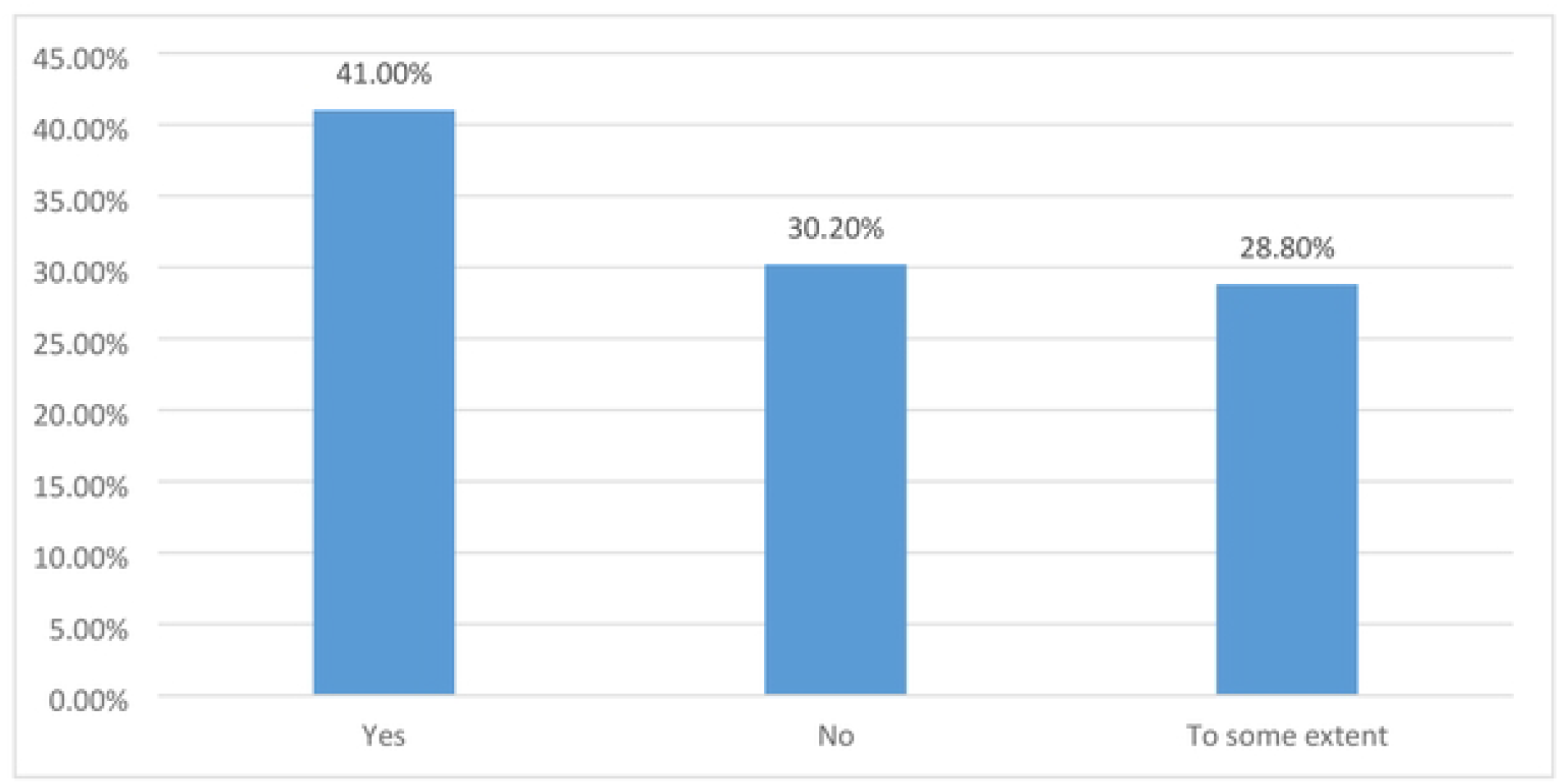
impact of current teaching methods on academic achievement among Omdurman Islamic University Medical Students-2021. Figure (15) shows that most of students (41%) reported that the university teaching methods had impact on their academic achievement, where 30.2% reported it had no impact and 28.8%reported that teaching methods had an impact to some extent.

The mean GPA of daily physically active students was within the range of 3.12 ± .51, weekly physically active students had a GPA mean of 3.06 ± .43 while the monthly physically active students had a GPA mean of 3.17 ± .47 with a P value of o.7, and this finding reveals that there is no significant relationship between frequency of PA and AA. (Table 4)

The mean GPA of students practice walking as main type of physical activity was within the range of 3.18 ± .46, students practice running as main physical activity had a GPA mean of 2.95 ± .57, students practice football as main physical activity had a GPA mean of 3.06 ± .47 while the other type of physical activity (gym - Zumba) had a

GPA mean of 3.04 ± .41 with a P value of o.2, this finding reveals, that there is no significant relationship between type of PA and AA.(Table 5)

Limitation of the study:

1. Small sample size.
2. Limiting study to one field.
3. Restricting study to one university and not accompanying others universities.

## 6.1 Conclusions

This study indicate that most students have sufficient information about physical activity, its importance and its impact on their academic and social life, and that most of them are physically active. The results also indicate that the main barrier for those who are not physically active (43.7%) is the courses load of the curriculum in the Faculty of Medicine, which requires dedication to most of the time for studying. Physical activity prefers (includes) was walking. The results also showed that there is no significant association between physical activity and academic achievement.

## 6.2 Recommendations

1. Recommendation to the students:
  a. Increase of physical activity and set aside a little time for it during the day.
  b. Allocate a good Space for physical activity through student activities and counseling their colleagues about its importance.
2. Recommendation to university leadership:
  a. Provide time for sports and physical activity and acknowledge its importance in the curriculum.
  b. Conduct regular sports programs under the supervision of the university to entertain students and stimulate their academic abilities.
  c. Encouraging and supporting research projects that studies the influential of academic achievement, particularly physical activity.
3. Recommendation to researchers:
  a. Conduct a study that includes a number of universities and a number of disciplines to measure the actual association between PA and AA.
  b. Conduct study with a large sample size to more representative and accurate

## Data Availability

we have no legal or ethical restrictions

## Notes

### Competing Interest Statement

The authors have declared no competing interest.

### Funding Statement

The author(s) received no specific funding for this work

## References and bibliography

1- Coleman KJ, Raynor HR, Mueller DM, Cerny FJ, Dorn JM, Epstein LH. Providing Sedentary Adults with Choices for Meeting Their Walking Goals. Preventive Medicine [Internet]. Elsevier BV; 1999 May;28(5):510–9. Available from: http://dx.doi.org/10.1006/pmed.1998.0471

2- Dishman RK, Heath G, Schmidt MD, Lee IM. Physical activity epidemiology. Human Kinetics; 2021.

3- Resaland GK, Moe VF, Aadland E, Steene-Johannessen J, Glosvik Ø, et al. Active Smarter Kids (ASK): Rationale and design of a cluster-randomized controlled trial investigating the effects of daily physical activity on children’s academic performance and risk factors for non-communicable diseases. BMC Public Health [Internet]. Springer Nature; 2015 Jul 28;15(1). Available from: http://dx.doi.org/10.1186/s12889-015-2049-y

4- Giles-Corti B, Donovan RJ. The relative influence of individual, social and physical environment determinants of physical activity. Social Science & Medicine [Internet]. Elsevier BV; 2002 Jun;54(12):1793–812. Available from: http://dx.doi.org/10.1016/S0277-9536(01)00150-2

5- Resaland GK, Aadland E, Moe VF, Aadland KN, Skrede T, Stavnsbo M, et al. Effects of physical activity on schoolchildren’s academic performance: The Active Smarter Kids (ASK) cluster-randomized controlled trial. Preventive Medicine [Internet]. Elsevier BV; 2016 Oct;91:322–8. Available from: http://dx.doi.org/10.1016/j.ypmed.2016.09.005

6- Melnyk BM, Jacobson D, Kelly S, Belyea M, Shaibi G, Small L, et al. Promoting Healthy Lifestyles in High School Adolescents. American Journal of Preventive Medicine [Internet]. Elsevier BV; 2013 Oct;45(4):407–15. Available from: http://dx.doi.org/10.1016/j.amepre.2013.05.013

7- 16- https://www.who.int/health-topics/physical-activity#tab=tab_1

8- Donnelly JE, Hillman CH, Castelli D, Etnier JL, Lee S, Tomporowski P, et al. Physical Activity, Fitness, Cognitive Function, and Academic Achievement in Children. Medicine & Science in Sports & Exercise [Internet]. Ovid Technologies (Wolters Kluwer Health); 2016 Jun;48(6):1197–222. Available from: http://dx.doi.org/10.1249/MSS.0000000000000901

9- Álvarez-Bueno C, Pesce C, Cavero-Redondo I, Sánchez-López M, Garrido-Miguel M, Martínez-Vizcaíno V. Academic Achievement and Physical Activity: A Metaanalysis. Pediatrics [Internet]. American Academy of Pediatrics (AAP); 2017 Nov 24;140(6):e20171498. Available from: http://dx.doi.org/10.1542/PEDS.2017-1498

10- https://www.definitions.net/definition/academic+achievement

11- 15- https://www.who.int/news-room/fact-sheets/detail/physical-activity

12- Steinmayr R, Meiǹer A, Weideinger AF, Wirthwein L. Academic achievement. Oxford University Press; 2014. DOI: 10.1093/obo/9780199756810-0108

13- Abdulghani HM, Al-Drees AA, Khalil MS, Ahmad F, Ponnamperuma GG, Amin Z. What factors determine academic achievement in high achieving undergraduate medical students? A qualitative study. Medical Teacher [Internet]. Informa UK Limited; 2014 Mar 11;36(Sup1):S43–S48. Available from: http://dx.doi.org/10.3109/0142159X.2014.886011

14- Niromand E, Salehi AR, Khazaei M, Khazaei MR. The Influential Factors in the Academic Achievement and Failure of Medical Students in Iran: A Review Study. Educational Research in Medical Sciences [Internet]. Kowsar Medical Institute; 2020 Dec 22;9(2). Available from: http://dx.doi.org/10.5812/ERMS.105860

15- Abdulghani HM, Al-Drees AA, Khalil MS, Ahmad F, Ponnamperuma GG, Amin Z. What factors determine academic achievement in high achieving undergraduate medical students? A qualitative study. Medical Teacher [Internet]. Informa UK Limited; 2014 Mar 11;36(Sup1):S43–S48. Available from: http://dx.doi.org/10.3109/0142159X.2014.886011

16- Ogundele GA, Olanipekun SS, Aina JK. Factors affecting students’ academic performance. Scholars Journal of Arts, Humanities and Social Sciences ISSN. 2014:2347–5374.

17- Khalil S, Almobarak AO, Awadalla H, Elmadhoun WM, Noor SK, Sulaiman AA, et al. Low levels of physical activity in Sudanese individuals with some features of metabolic syndrome: Population based study. Diabetes & Metabolic Syndrome: Clinical Research & Reviews [Internet]. Elsevier BV; 2017 Dec;11:S551–S554. Available from: http://dx.doi.org/10.1016/j.dsx.2017.04.003

18- Abu Saa A, Al-Emran M, Shaalan K. Factors Affecting Students’ Performance in Higher Education: A Systematic Review of Predictive Data Mining Techniques. Technology, Knowledge and Learning [Internet]. Springer Science and Business Media LLC; 2019 Apr 25;24(4):567–98. Available from: http://dx.doi.org/10.1007/S10758-019-09408-7

19- Al-Drees A, Abdulghani H, Irshad M, Baqays AA, Al-Zhrani AA, Alshammari SA, et al. Physical activity and academic achievement among the medical students: A cross-sectional study. Medical Teacher [Internet]. Informa UK Limited; 2016 Mar 17;38(sup1):S66–S72. Available from: http://dx.doi.org/10.3109/0142159X.2016.1142516

20- Chung Q-E, Abdulrahman SA, Jamal Khan MK, Jahubar Sathik HB, Rashid A, et al. The Relationship between Levels of Physical Activity and Academic Achievement among Medical and Health Sciences Students at Cyberjaya University College of Medical Sciences. Malaysian Journal of Medical Sciences [Internet]. Penerbit Universiti Sains Malaysia; 2018;25(5):88–102. Available from: http://dx.doi.org/10.21315/MJMS2018.25.5.9

21- Fenuta A, Brennan A, Lau R, Shirazipour C, Hefnawi B, D’Urzo K, Johnson A, McPhee I, McEachern B, Tomasone J. Medical Student Physical Activity Education– Staying Active to Actively Help Others. MedEdPublish. 2019 Oct 22;8.

